# Examining the possible causal relationship between Lung Function, COPD and Alzheimer’s Disease. A Mendelian Randomization Study

**DOI:** 10.1101/2020.08.27.20182964

**Authors:** Daniel H Higbee, Raquel Granell, Esther Walton, Roxanna Korologou-Linden, George Davey Smith, James W Dodd

**Author notes:** Corresponding Author: Dr James Dodd. All authors made substantial contributions to analysis, drafting and final approval of the paper. DHH, JWD, RG and GDS were involved in the conception.

## Abstract

**Rationale:** Large retrospective case-control studies have reported an association between COPD, reduced lung function and an increased risk of Alzheimer’s disease. However, it remains unclear if these diseases are causally linked, or due to shared risk factors. Conventional observational epidemiology suffers from unmeasured confounding and reverse causation. Additional analyses addressing causality are required.

**Objectives:** To examine a causal relationship between COPD, lung function and Alzheimer’s disease.

**Methods:** Using two-sample Mendelian randomization, we utilised single nucleotide polymorphisms (SNPs) identified in a genome wide association study (GWAS) for lung function as instrumental variables (exposure). Additionally, we used SNPs discovered in a GWAS for COPD in those with moderate to very severe obstruction. The effect of these SNPs on Alzheimer’s disease (outcome) were taken from a GWAS based on a sample of 24,807 patients and 55,058 controls.

**Results:** We found minimal evidence for an effect of either lung function (odds ratio [OR]:1.02 per SD; 95% confidence interval [CI]: 0.91-1.13; p-value 0.68). or liability for COPD on Alzheimer’s disease (OR: 0.97 per SD; 95% CI: 0.92 – 1.03; p-value 0.40).

**Conclusion:** Neither reduced lung function nor liability COPD are likely to be causally associated with an increased risk of Alzheimer’s, any observed association is likely due to unmeasured confounding. Scientific attention and health prevention policy may be better focused on overlapping risk factors, rather than attempts to reduce risk of Alzheimer’s disease by targeting impaired lung function or COPD directly.

## Introduction

Chronic Obstructive Pulmonary Disease (COPD) is a disease of multi-morbidity^(1)^. In COPD the presence of multi-morbidity is associated with higher mortality, worse quality of life and increased healthcare utilisation ^(2, 3)^. Impaired lung function measures such as Forced Expiratory Volume in one second (FEV_1_) and Forced Expiratory Volume (FVC) have been found to be strongly associated with multi-morbidity.^(4)^ However, it remains unclear if these multi-morbidities are causally linked to lung function and disease, for example through a proposed inflammatory overspill, or if they are due to shared risk factors, such as smoking ^(5)^. Therapeutic targets may be identified if specific causal mechanisms could be established.

Cognitive impairment is a common co-morbidity in COPD, with reported prevalence ranging from 10-61% and around 25% of older adults with dementia also have COPD^(6)^. Cognitive impairment in COPD is associated with greater disability^(7)^, poorer medication compliance,^(8)^ and risk of exacerbation and mortality^(7)^. Poor pulmonary function in early life has been associated with increased odds of dementia later in life, even after adjustment for smoking.^(9)^

Alzheimer’s disease (AD) is the most common type of dementia^(10)^, its association with COPD is less well defined than general cognitive ability, but reports of a potential link between COPD and AD was first described nearly 30 years ago^(11)^. Large retrospective observational case-control cohorts have reported increased risk of AD in patients with both COPD and reduced lung function^(12, 13)^. For example, Lutsey et al reviewed hospitalisation codes^(13)^ in the Atherosclerosis Risk In Communities Study for AD-related outcomes and reported that an Odds Ratio of 1.24 for AD-type dementia or mild cognitive impairment (MCI) in patients with COPD and OR 1.79 for those with a restrictive impairment compared to controls. If the lung function and COPD have a causal effect on risk of Alzheimer’s disease, then they could be modifiable risk factors.

Mendelian Randomization (MR) is an established genetic epidemiological method which can overcome problems of unmeasured confounders and reverse causation, typical of conventional observational epidemiology^(14)^. MR allows causal inference through the use of genetic variants as proxies for non-genetic (modifiable) risk factors or health outcomes^(14)^. MR uses genetic data, e.g. single nucleotide polymorphisms (SNPs) that are associated with an exposure (in this case diagnosis of COPD or lung function), and uses them as instrumental variables (IV) to assess the causal effect of the exposure on the outcome of interest (in this case Alzheimer’s disease)^(15)^.

MR has multiple advantages. Genetic variants are randomly allocated at birth which can be exploited to simulate randomisation^(15)^. They are not influenced by behavioural or environmental factors minimizing reverse causality (where the outcome, or early stages of the disease process that leads to the outcome, influences the exposure) ^(20)^. Additionally, the effects are equivalent to lifetime differences, reducing issues relating to transient fluctuations. Our objective was to use MR to investigate if there is any evidence of a causal effect between the exposures, lung function and liability to COPD and the outcome, Alzheimer’s disease.

## Methods

### Lung Function

We used data from Shrine et al. the largest currently available lung function GWAS, n=400,102 ^(16)^ which reported 279 genome wide significant SNPs (p<5*10^-9^). Lung function measurements used were Forced Expiratory Volume in 1 second (FEV_1_), Forced Vital Capacity (FVC), FEV_1_/FVC ratio and Peak Expiratory Flow (PEF). A weighted risk score, was associated with risk of COPD (p= 6.64×10^-63^), with an Odds ratio of 1.55 for each standard deviation of the risk score ^(16, 17)^. Further details of the study population can be found in the supplementary information and the reference ^(16)^.

### COPD

We used 82 SNPs associated with COPD, as identified in Sarkonsakaplat et al. case control GWAS^(18)^, n = 35,735 cases and 222,076 controls discovered in meta-analysis of 25 studies. COPD was defined by Global Initiative for Chronic Obstructive Lung Disease criteria; FEV_1_/FVC <0.7 and FEV_1_ <80% predicted. Further details of study population can be found in the supplementary information and the reference.^(18)^

80% and 77% of the Shrine et al and Sarkonsakaplat et al GWAS sample respectively were from the UKBiobank ^(19)^. In brief the UK Biobank is a large prospective cohort study where >500,000 participants were recruited from 2006 – 2010 in the United Kingdom (54% female). Pre-bronchodilation lung function testing was performed by trained healthcare staff.

### Alzheimer’s disease

We used data from a meta-analysis of the International Genomics of Alzheimer’s disease (IGAP) consortium^(20)^, Alzheimer’s Disease Sequencing Project (ADSP) ^(21)^, and Psychiatric Genomics Consortium (PGC) ^(22)^ totalling 24,807 Alzheimer’s disease cases and 55,058 controls ^(23)^. All cases had clinical diagnoses of Alzheimer’s disease. Some participants of the ADSP cohort were previously also included in IGAP, so ADSP individuals that were duplicates based on the comparison of individual level genetic data between IGAP and ADSP were excluded.

There was no sample overlap between the exposure and outcome samples. All participants were of European ancestry.

### Statistical Analysis

Statistical analysis was done using R Studio version 3.5.1. and the MRCIEU/TwoSampleMR R package ^(24)^.

For exposure traits r^2^ was estimated for each genetic instrument and used to determine the Cragg-Donald overall F-statistic.^(25)^ The higher the F-statistic the lower the chance of weak instrument bias.^(25)^ For all exposures SNPs LD-clumping was performed using European reference population (kb = 10000, r^2^ 0.001). Palindromic SNPs (i.e. A/T and C/G SNPs) with intermediate allele frequencies were excluded. The remaining SNPs were harmonised^(26)^. Steiger filtering^(27)^ was performed to remove variants that caused more variance of the outcome than the exposure, see online supplement for more details.

### Main Mendelian Randomization Analysis

Inverse Variance Weighting (IVW) was used for main effect estimate. This is a weighted regression of SNP-outcome on SNP-exposure associations combined where the y intercept is constrained to zero.

### Assumption and Sensitivity Analysis

MR assumptions and further details of tests used, are detailed in the online appendix. To account for the possibility of horizontal pleiotropy (IVs influence exposure and outcome through independent pathways), we performed MR Egger. To minimise the effect of unbalanced instruments on an overall estimate of the mean, weighted median and mode MR methods were performed. To assess for horizontal pleiotropy a funnel plot was made by plotting the effect against its precision (beta against standard error). To ensure the results were not due to outliers with a large effect, a leave-one-out analysis was performed by re-estimating the total effect after sequentially excluding one SNP at a time and a single-SNP analysis, where the effect of each SNP was individually assessed via IVW analysis and represented in a forest plot.

Heterogeneity (the variability in causal estimates obtained for each SNP) is an indication of potential violation of assumptions. This was calculated and assessed with a Q statistic.

## Results

F-statistic for lung function GWAS^(16)^ exposures were; All traits= 111, FEV_1_=69, FVC=70, FEV_1_/FVC=148, making weak instrument bias unlikely. After clumping, extracting SNPs from outcome GWAS, Steiger filtering and removal of palindromic SNPs, 131 SNPs were available for analysis. See flow charts in supplementary information for detailed analysis pathway.

We found minimal evidence for a causal effect of lung function (all traits) on Alzheimer’s disease, (IVW odds ratio [OR]:1.02 per SD; 95% CI: 0.91-1.13; p-value 0.68). This result was further confirmed in a sensitivity analysis using both weighted median (OR: 1.01 per SD; 95% CI:0.86-1.19, p value = 0.81), and weighted mode MR (OR 0.99 per SD;95% CI 0.78-1.19), p-value = 0.81). risk of AD. The MR-Egger causal estimation produced similar results with an OR 1.05 per SD (95% CI 0.79-1.34; p-value 0.71). The confidence interval of the MR-Egger is wider than that of IVW, consistent with the lower statistical power of this test.

**Figure 1** plots each individual SNP-exposure effect against SNP-outcome with the coloured lines representing each statistical test. Increasing lung function (exposure) does not have a consistent effect on Alzheimer’s disease (outcome).

**Figure 1.**
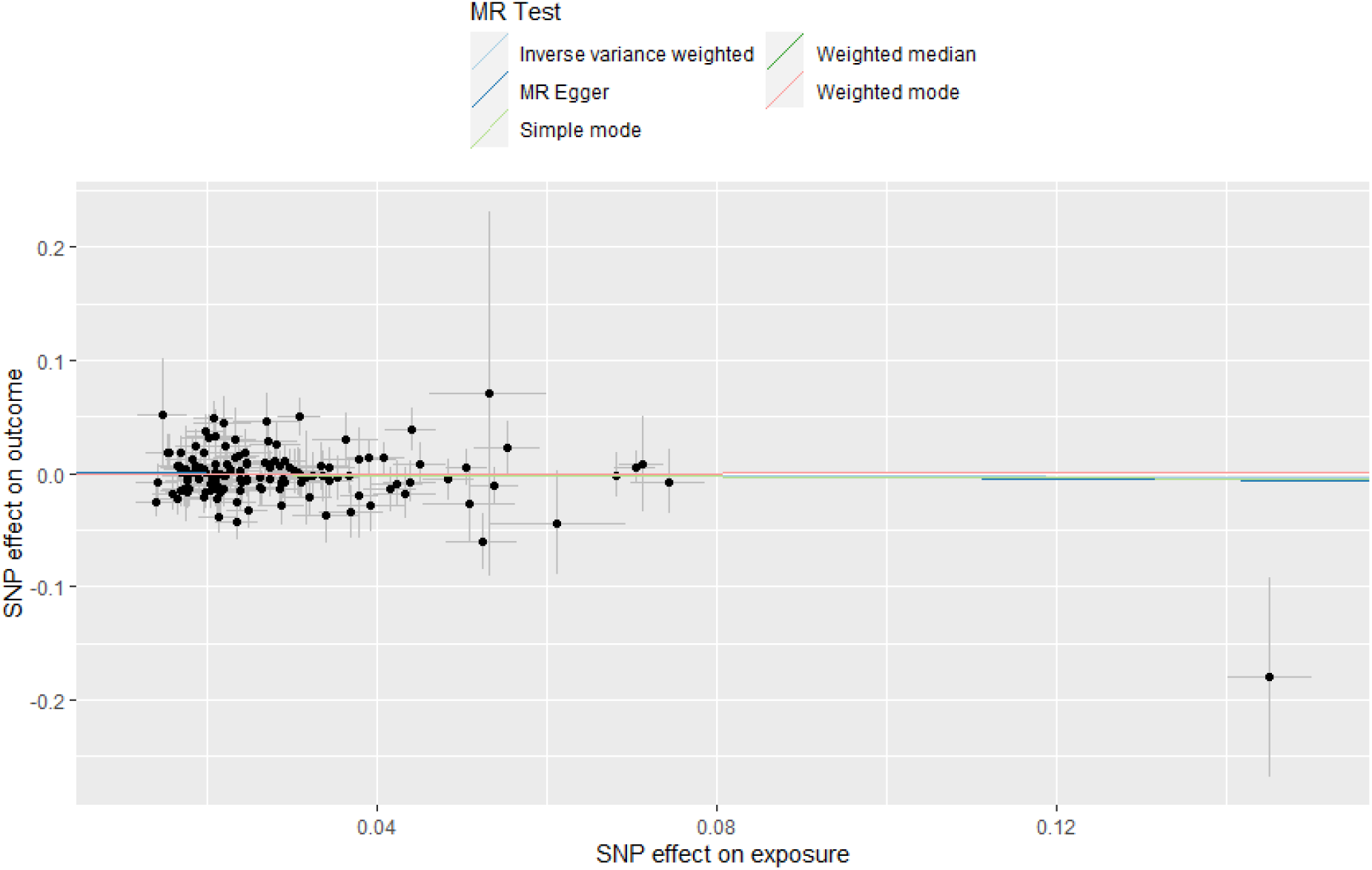
Scatter plot of the SNP-effect on lung function trait and SNP-effect on Alzheimer’s disease. Each point on the graph represents the SNP-outcome association plotted against the SNP-exposure association. Bars indicate 95% confidence intervals. Coloured lines represent analysis method used. This shows no effect of lung function on Alzheimer’s disease. MR Egger intercept is close to zero indicating no unbalanced directional pleiotropy.

**Table 1** shows that these results were consistent when analysing lung function traits FEV_1_, FVC, and FEV_1_/FVC individually with little evidence of a causal association on Alzheimer’s disease with confidence intervals crossing 1 for all statistical tests.

**Table 1.** Two-sample MR results of lung function traits^(16)^ on Alzheimer’s disease^(23)^

|  |  | Lung Function Trait (Exposure) |  |  |  |
| --- | --- | --- | --- | --- | --- |
|  |  | FEV<sub>1</sub>, FVC,<br>FEV<sub>1</sub>/FVC, PEF | FEV<sub>1</sub> | FVC | FEV<sub>1</sub>/FVC |
| No.<br>SNPs<br>used |  | 131 | 42 | 46 | 73 |
| IVW | OR per SD | 1.02 | 1.04 | 1.08 | 0.99 |
|  | (95% CI) | (0.91 – 1.13) | (0.82 – 1.32) | (0.85 – 1.37) | (0.88 – 1.13) |
|  | p-Value | 0.68 | 0.73 | 0.51 | 0.97 |
|  | Q_p-value* | 0.26 | 0.30 | 0.19 | 0.71 |
| Weighted<br>Median | OR per SD | 1.01 | 1.15 | 1.14 | 0.95 |
|  | (95% CI) | (0.86 – 1.19) | (0.82 – 1.61) | (0.83 – 1.58) | (0.79 – 1.15) |
|  | p-Value | 0.81 | 0.39 | 0.39 | 0.62 |
| Weighted<br>Mode | OR per SD | 0.99 | 1.07 | 1.04 | 0.97 |
|  | (95% CI) | (0.78 – 1.26) | (0.60 - 1.90) | (0.61 – 1.78) | (0.74 – 1.26) |
|  | p-Value | 0.97 | 0.80 | 0.86 | 0.84 |
| MR-Egger | OR per SD | 1.05 | 1.22 | 0.97 | 0.95 |
|  | (95% CI) | (0.79 – 1.34) | (0.57 - 2.59) | (0.36 – 2.62) | (0.69 – 1.31) |
|  | p-Value | 0.71 | 0.59 | 0.96 | 0.77 |

\*A test for heterogeneity. If this was <0.05 it would suggest heterogeneity
OR – Odd ratio; CI – Confidence Interval; IVW – Inverse Variance Weighting

We used single-SNP analyses to determine the effect of each lung function SNP on the odds of Alzheimer’s disease **(Figure 2)**. The SNP rs2070600, may be an outlier due to its comparatively large effect on both lung function and AD. Polymorphisms in this SNP have been described as having a weak effect on Alzheimer’s disease risk.^(28)^ However, despite excluding this SNP from the analysis the results were similar (e.g. see leave-one-out analysis in **Figure 3**).

**Figure 2.**
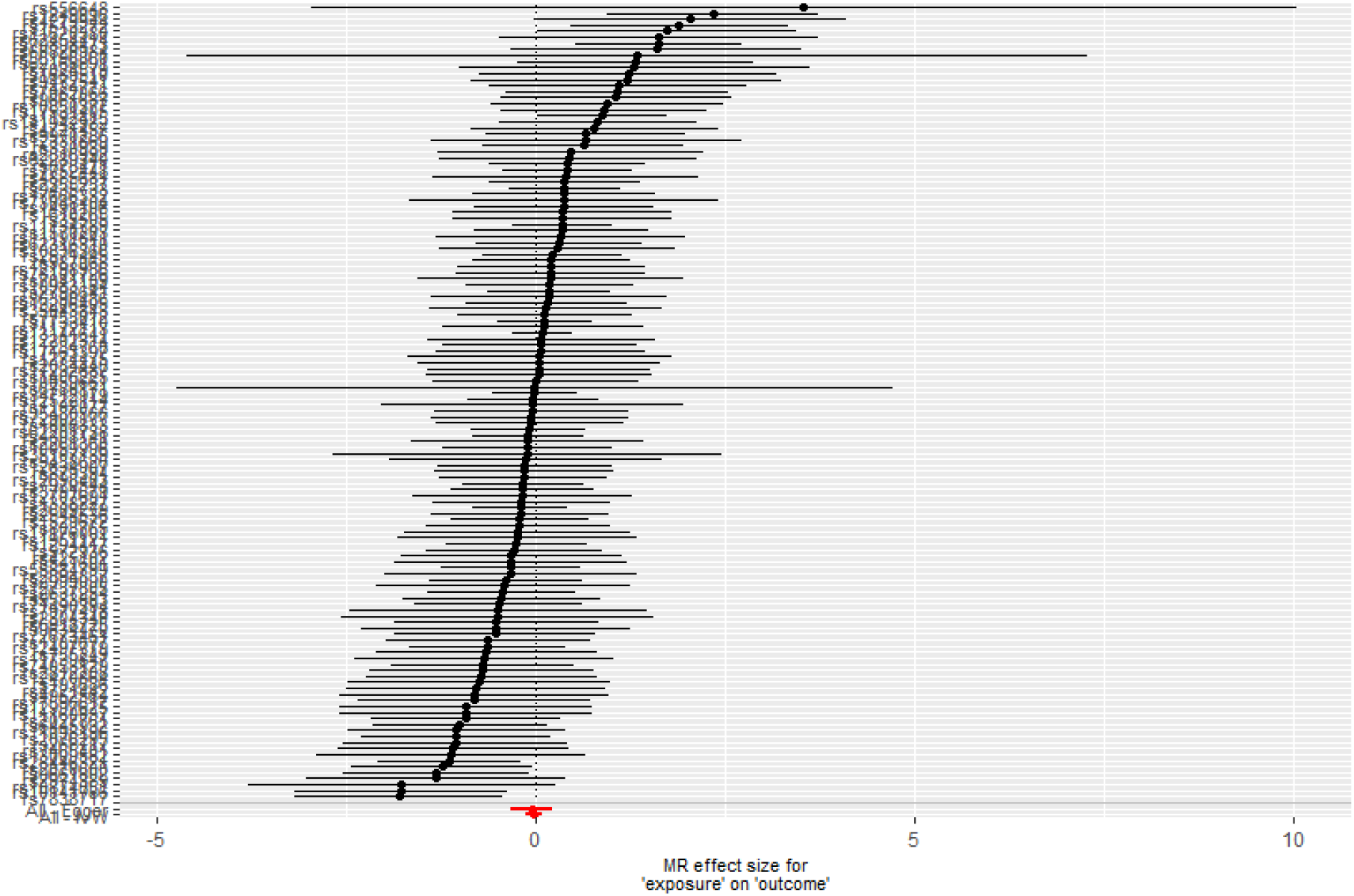
Single SNP analysis of lung function traits on Alzheimer’s disease. Each point represents individual SNP calculated effect size for lung function on the odds of Alzheimer’s disease. Bars indicate 95% CI.

**Figure 3.**
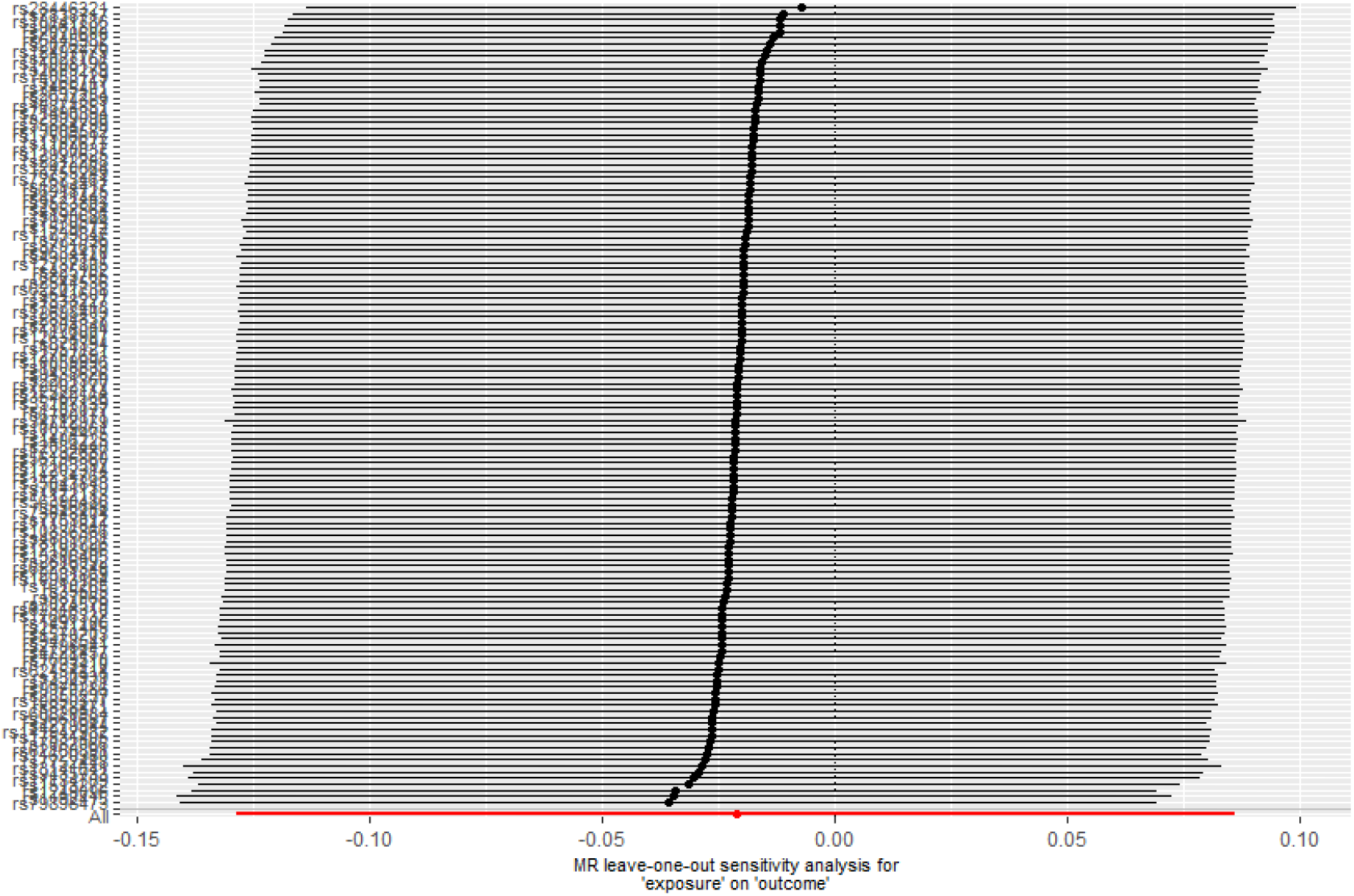
Leave-one-out analysis of lung function traits on Alzheimer’s disease. Each point represents the IVW estimate if the SNP on the y axis was left out of total analysis. Bars indicate 95% confidence intervals, demostrating that no individual SNP is driving the causal effect estimate.

Each SNP beta was plotted against its inverse standard error (**Figure 4)** producing a funnel shape indicating no heterogeneity. In addition to these visual tests, we found little evidence of heterogeneity using a Q statistic when lung function traits were combined or assessed individually (**Table 1**. Q_p value >0.51). MR-Egger intercept was <0.001, visually displayed in **Figure 1**, indicating there was no unbalanced horizontal pleiotropy.

**Figure 4.**
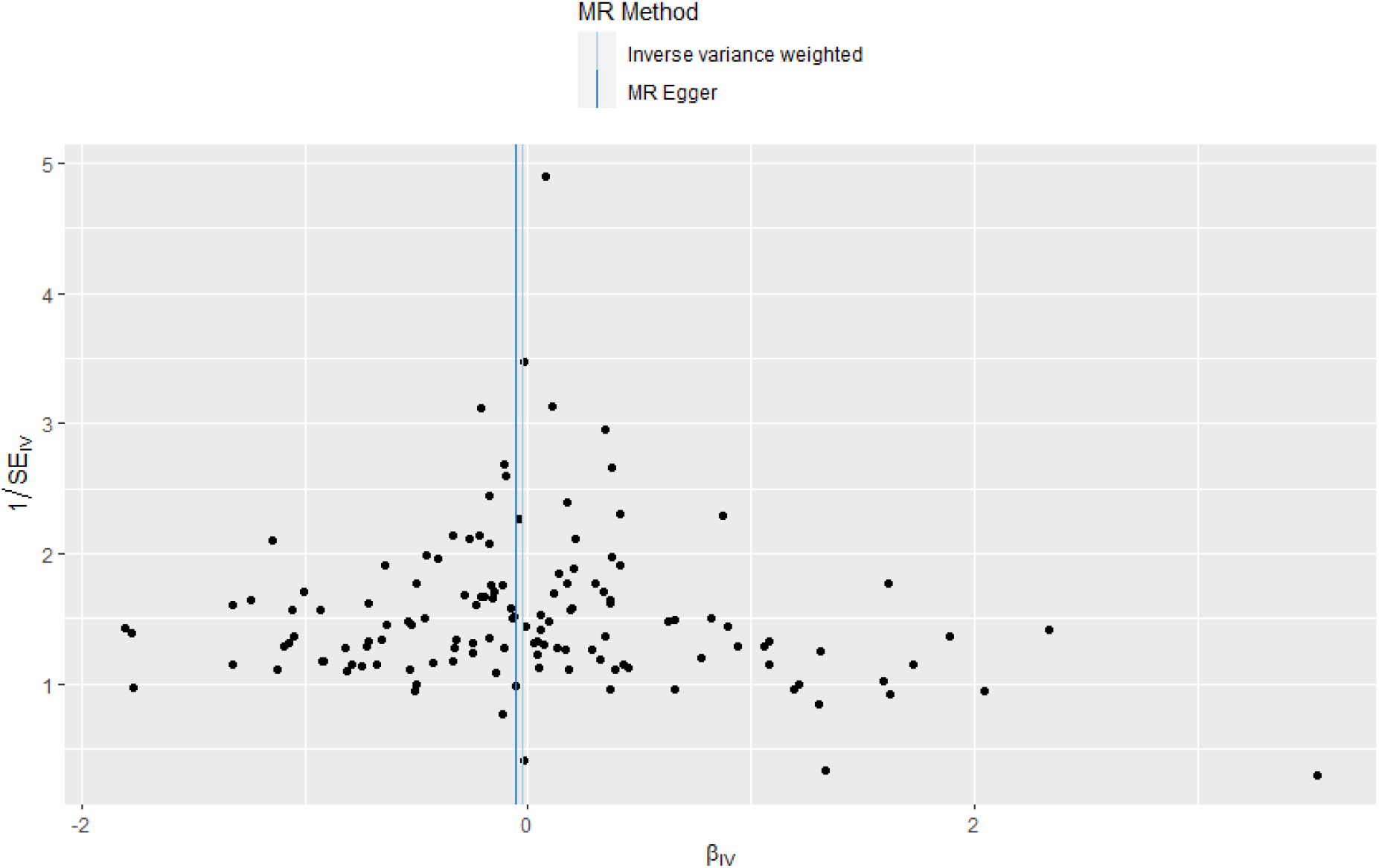
Funnel plot of heterogeneity of causal effects of lung function traits on Alzheimer’s disease. Each point is a SNP with its beta plotted against its inverse standard error. As the graph is funnel shaped, it indicates no heterogeneity.

Using the COPD GWAS^(18)^ gave an F-statistic of 52. After clumping, extracting SNPs from outcome GWAS, Steiger filtering and removal of palindromic SNPs, 53 SNPs for liability to COPD were available for analysis in the Alzheimer’s outcome GWAS. Please see flow chart in supplementary information for details. Results are displayed in **Table 2**.

**Table 2.** Two-sample MR results of COPD^(18)^ on Alzheimer’s disease^(29)^

| COPD |  |  |
| --- | --- | --- |
| No. SNPs used |  | 53 |
| IVW | OR per SD (95% CI) | 0.97 (0.92 – 1.03) |
|  | p-Value | 0.40 |
| | $Q_p$-value | 0.57 |
| Weighted Median | OR per SD (95% CI) | 0.97 (0.90 – 1.05) |
|  | p-Value | 0.52 |
| Weighted Mode | OR per SD (95% CI) | 0.96 (0.86 – 1.08) |
|  | p-Value | 0.56 |
| MR-Egger | OR per SD (95% CI) | 1.10 (0.93 – 1.31) |
|  | p-Value | 0.23 |

OR – Odd ratio; CI – Confidence Interval; IVW – Inverse Variance Weighting

We found minimal evidence for an effect of liability to COPD on risk of Alzheimer’s disease (IVW OR: 0.97 per SD; 95% CI: 0.92 – 1.03; p-value 0.40). This result was further confirmed in our sensitivity analysis using both weighted median (OR: 0.97 per SD; 95% CI: 0.90-1.05; p-value = 0.52), and weighted mode MR (OR: 0.96 per SD; 95% CI: 0.86-1.08; p-value = 0.56). The MR-Egger causal estimation produced an OR 1.11 per SD (95% CI: 0.93-1.31; p-value 0.2), the only test to show a direction of effect of increasing COPD causing increased risk of Alzheimer’s disease. **Figures 5-8** are available in supplementary information, demonstrating that results were not driven by an individual SNP. There was no evidence of heterogeneity, with a Q-pvalue 0.57.

## Discussion

### Evidence before this study

Our results indicate that there is minimal evidence of a causal association between lung function or liability to COPD and risk of Alzheimer’s disease. This is in contrast to two large observational studies^(12, 13)^, which do report an association between COPD and Alzheimer’s disease. The observed associations may be due to unmeasured confounding by risk factors common to both COPD and Alzheimer’s disease such as smoking, physical inactivity, social deprivation and lower educational attainment^(30)^. The observational studies may have inadvertently included other forms of dementia other than Alzheimer’s disease, for example vascular dementia resulting from cerebrovascular or neurological damage. Apolipoprotein e4 allele is the biggest risk factor for Alzheimer’s disease whereas it is thought that COPD affects cognition via vascular effects. There is evidence that COPD and reduced lung function is associated with micro and macrovascular damage that could mediate the relationship.^(31-33)^ It is possible that vascular dementia is causally linked to COPD and lung function, but this outcome was not included in our analysis which was restricted to Alzheimer’s disease only. Cognitive dysfunction and Mild Cognitive Impairment are well described in COPD ^(6)^. It may be that this association is causal, but that patients do not progress to Alzheimer’s disease due to their lung disease. Survivor bias (where selection is conditional upon survival to recruitment^(34)^) can be of concern in studies involving potentially fatal diseases of later life. Potentially, patients with COPD would be less likely to be recruited to a GWAS, biasing the MR towards a null. Observational studies performed by analysing health records may be less likely to be affected by this.

### Impact of this study

This analysis uses two-sample MR and multivariable MR to explore a causal association between lung function, COPD and Alzheimer’s disease. The increasing incidence of Alzheimer’s disease in Western society has been described as an epidemic ^(35)^. COPD is responsible for 5% of global disability-adjusted life years and 5% of total deaths ^(36)^. Consequently, prevention and treatment of both COPD and Alzheimer’s disease is a global health priority. Although there have been efforts to search for causal mechanisms linking the two diseases, our analysis using multiple means of assessing causation would suggest scientific attention and health prevention resources may be better focused on overlapping risk factors such as smoking, diet and physical activity ^(37, 38)^, rather than attempts to reduce risk of AD by improving lung function or reducing liability to COPD alone.

### Strengths and Limitations

By using randomly assigned genetic variants as an exposure, two-sample MR methodology eliminates many confounders in observational epidemiology^(14)^. We used a large number of robust lung function SNPs, which have been well validated in large samples.^(16, 18)^ It is important to ensure that the assumptions of MR are met when dealing with SNPs for complex phenotypes like lung function and COPD. We adhered to proposed methodological guidelines of MR (STROBE)^(39)^ which are designed to increase reliability of MR reporting. None of the sensitivity tests provided strong evidence for a violation of the MR assumptions. COPD is a binary trait, so our SNPs confer liability to COPD. As this is a Two Sample MR study, we do not know how many participants in the outcome population had COPD. COPD is a clinical diagnosis with set spirometric thresholds, whereas in the discovery GWAS a diagnosis of COPD was made based on spirometric criteria alone. This was done by dichotomising continuous traits. Dichotomization of continuous traits in MR studies can make interpretation of the causal estimate less reliable, but MR can still be a valid test of the causal null hypothesis for a binary exposure.^(40)^

As the SNPs were discovered in populations of those with European ancestry, the results may not be generalisable to other populations. However, we believe the findings of this study are likely to be generalisable to the non-European population as a genetic risk score of the lung function SNPs was validated in other ancestral populations, with slightly reduced effect ^(16)^.

### Conclusions

Lung function and liability to COPD are not causally associated with an increased risk of Alzheimer’s disease. Previous observational studies showing and association between impaired lung function or COPD and Alzheimer’s disease are most likely due to unmeasured confounding.

## Data Availability

Code available on request from corresponding author. Data used was summary data freely available in supplementary tables or from corresponding authors of discovery GWAS.

## Acknowledgements

We acknowledge participants and investigators of the IGAP^(20)^, ADSP^(21)^, PGC^(22)^, UKBiobank and Spirometa groups, and the authors of the Alzheimer’s disease^(23)^ lung function^(16)^ and COPD^(18)^ GWAS.

## Online Data Supplement

Contents

Appendix 1. Figures for COPD specific SNPs

Appendix 2. Assumptions and tests

Appendix 3. Details of sample populations

Appendix 4. Flow chart of analyses

Appendix 5. Data and code availability

## Appendix 1. Figures for COPD SNPs

**Figure E1.**
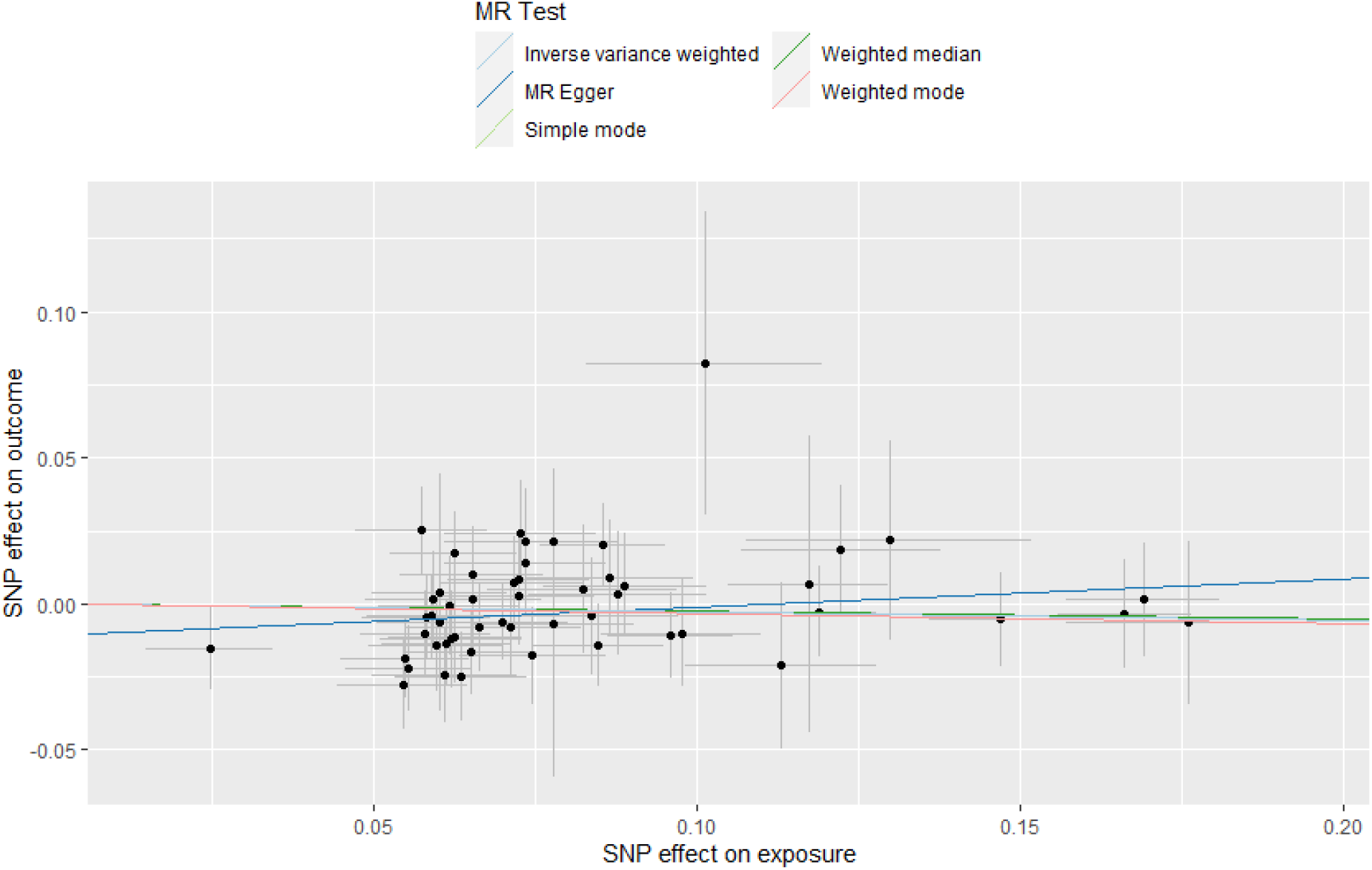
Scatter Plot of IVW COPD and AD. Each point on the graph represents the SNP-outcome association plotted against the SNP-exposure association. Bars indicate 95% confidence intervals. Coloured lines represent analysis method used. This shows no signficant effect of COPD on Alzheimer’s disease. MR Egger intercept is close to zero indicating no unbalanced directional pleiotropy.

**Figure E2.**
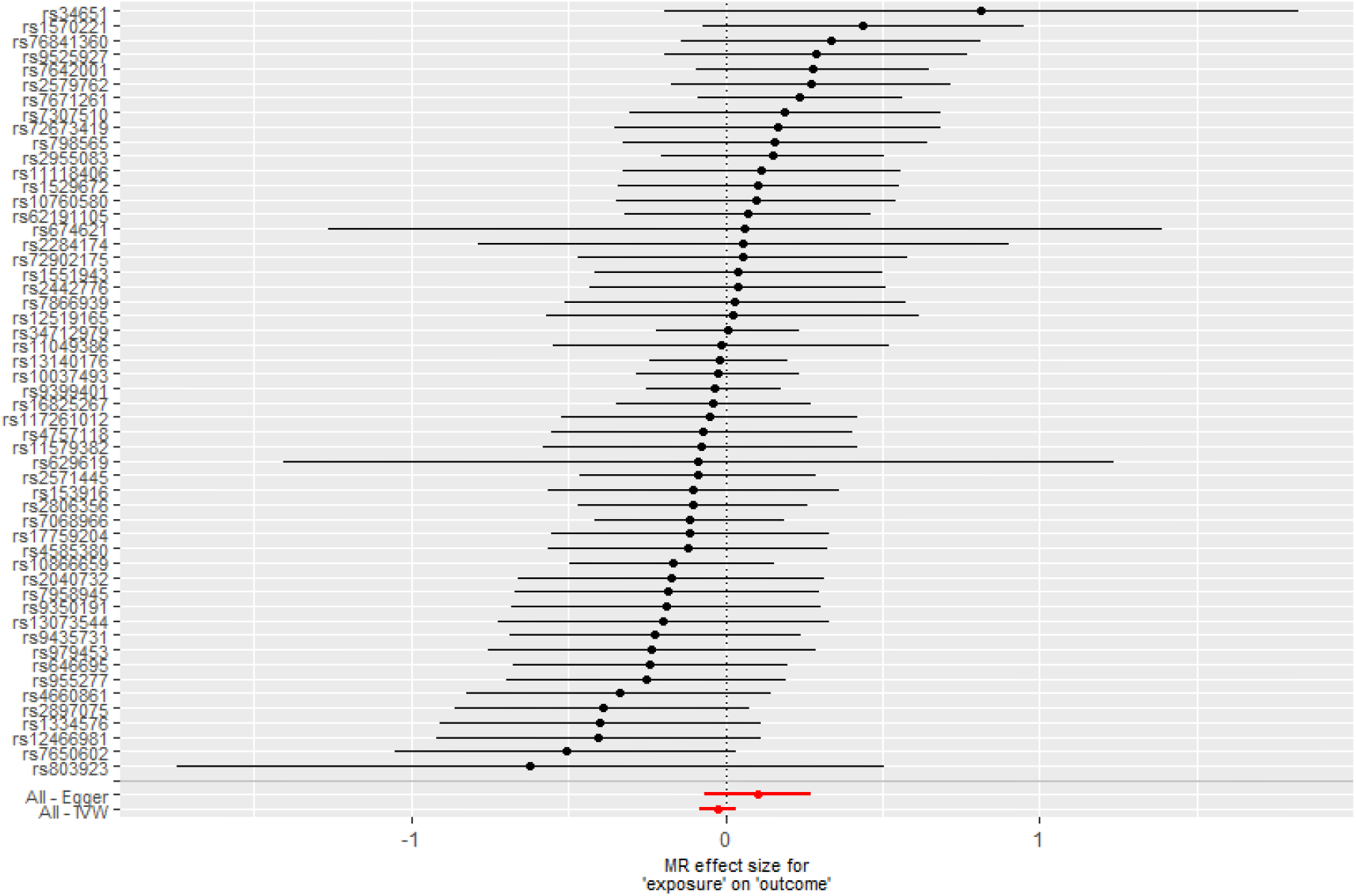
Single SNP Analysis of COPD and AD. Each point represents individual SNP calculated effect size for COPD on odds of Alzheimer’s disease. Bars indicate 95% CI.

**Figure E3.**
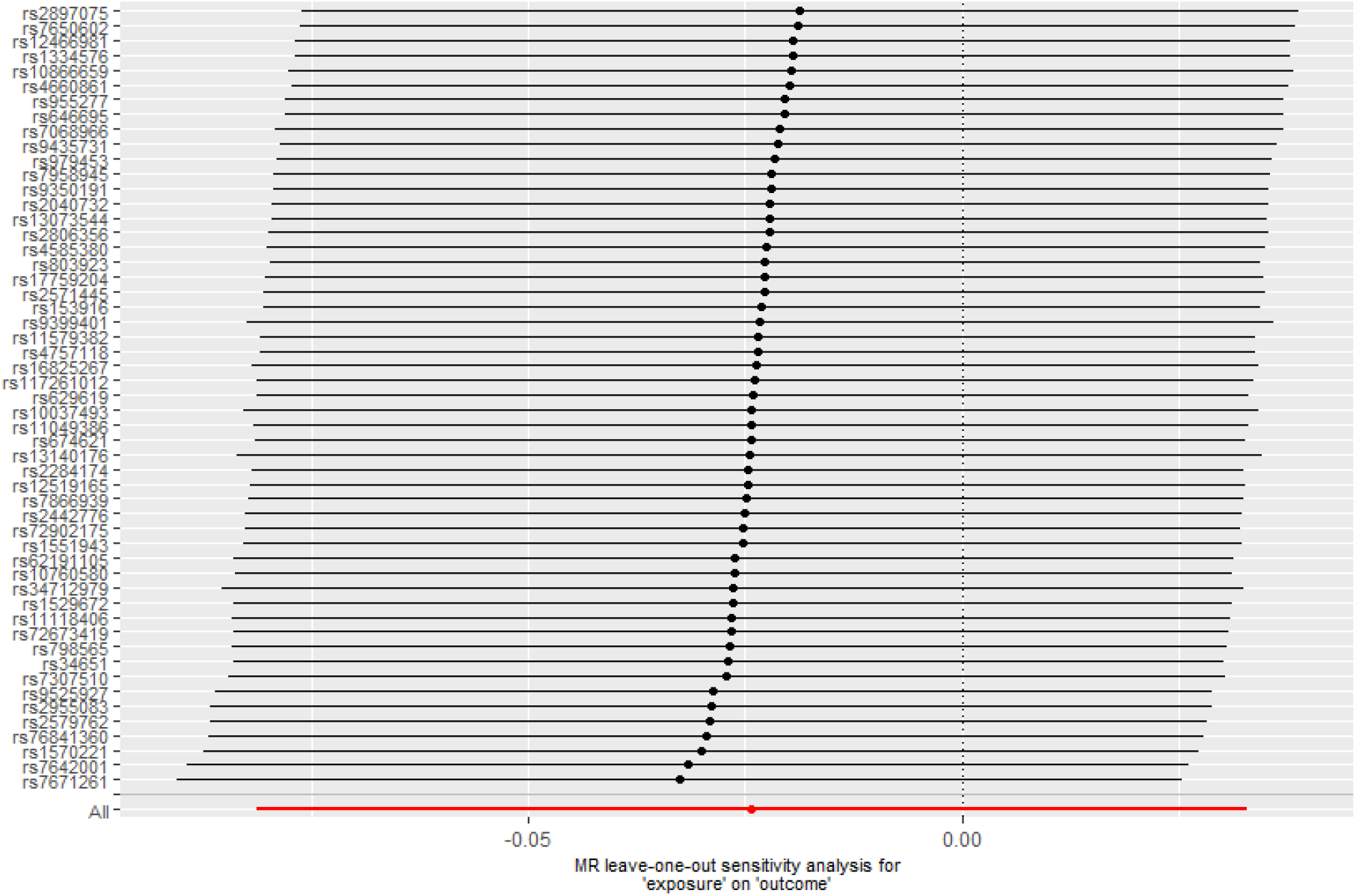
Leave one out analysis of COPD and AD. Each point represents the IVW estimate if the SNP on the y axis was left out of total analysis. Bars indicate 95% confidence intervals. It demostrates that no individual SNP is driving the causal effect estimate.

**Figure E4.**
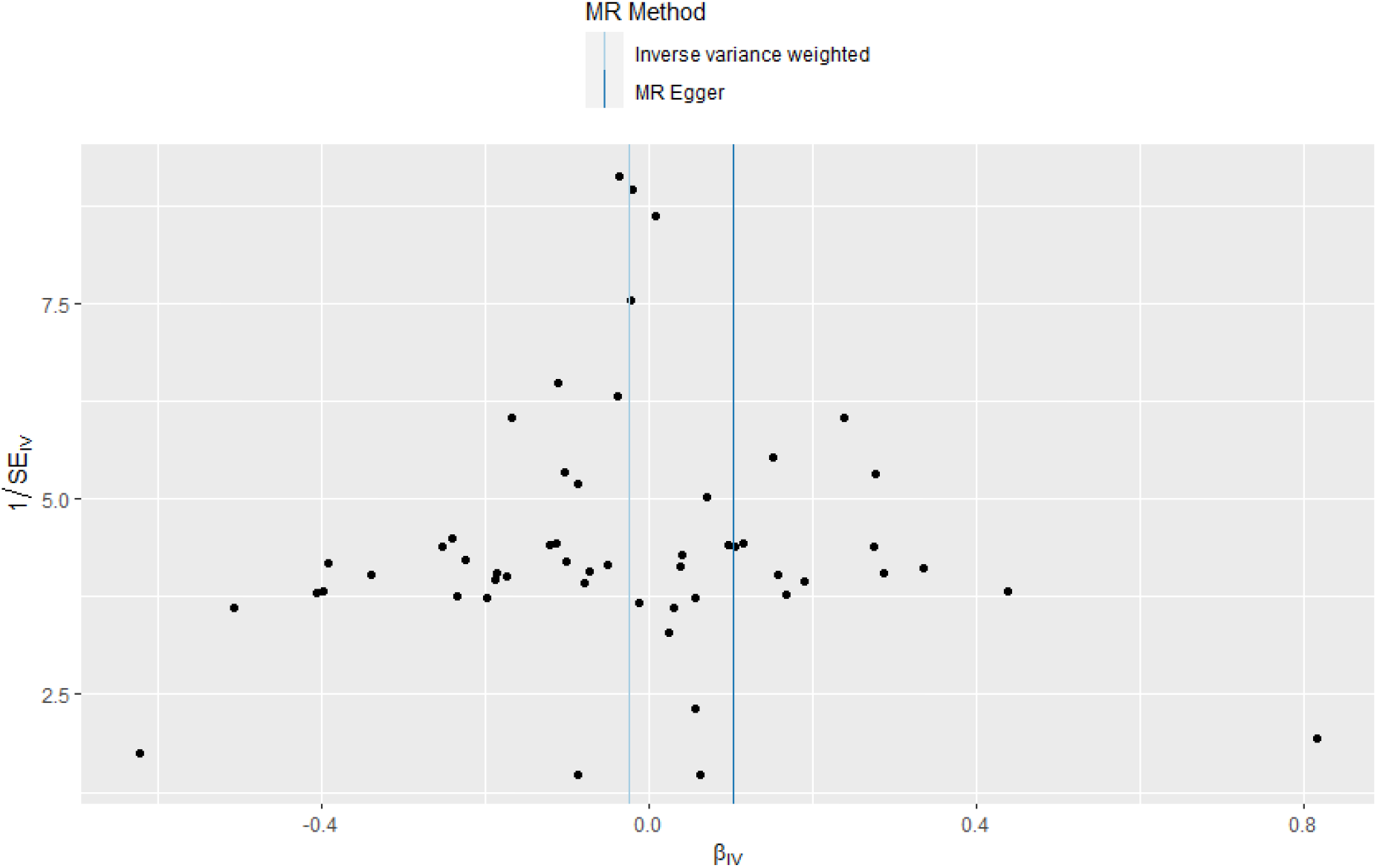
Funnel Plot analysis of COPD and AD. Each point is a SNP with its beta plotted against its inverse standard error. As the graph is funnel shaped, it indicates no heterogeneity.

## Appendix 2

### Assumptions

We assume that our IVs have a true association with the exposures. This has been rigorously statistically tested in the discovery GWAS and effect estimation ^(1, 2)^. F-statistic calculation shows that all exposure SNPs were unlikely to be weak instruments. In both GWAS papers SNPs discovered are related to specific genes, cell types and biological pathways for lung tissue development.

We assume that our SNPs do not affect AD except via their effect on LF/COPD, and that the SNPs have no associations with any confounders that are also associated with AD. Although not possible to directly tests, our sensitivity and heterogeneity tests reduce the risk these assumptions were violated. To account for the possibility of horizontal pleiotropy (IVs affect multiple pathways) we performed MR Egger, weighted median and weighted mode tests. MR-Egger is similar to IVW except the y intercept is unconstrained. If the y intercept of the MR-Egger is not equal to zero then either there is unbalanced horizontal pleiotropy (the average pleiotropic effect differs from zero) or the pleiotropic effects are independent from the genetic association with the risk factor, or both ^(3)^. Although power is lower compared to IVW, the gradient of the MR-Egger gives a causal estimate of the dose-response relationship between the genetic associations with the risk factor and those with the outcome, providing additional evidence for causal affect. To help avoid the effect of unbalanced instruments on an overall estimate of the mean by the IVW method, weighted median and mode MR methods were performed. A weighted median MR gives a consistent estimate of the causal effect when at least 50% of the weight comes from valid IVs, giving a greater robustness with strongly outlying causal estimates ^(4)^. A weighted mode MR calculates an estimate based on the set of SNPs that form the largest homogenous cluster, which attempts to avoid the impact of invalid instruments ^(5)^.

There was no evidence of population stratification (when subgroups within a sample are of different genetic ancestry) as assessed by linkage disequilibrium score regression in the original GWAS.

### Steiger Filtering

Steiger filtering estimates each SNP’s rsq.exposure and rsq.outcome in the outcome population.^(6)^ Those SNPs that explain more variance in the outcome than exposure are excluded, as they could led to a reverse causal relationship. SNPs were removed if they explained more variance of the outcome than the exposure. SNPs were removed if they explained more variance of the outcome than the exposure. Necessary information to perform Steiger filtering includes knowing the case and control numbers for each SNP. In our main Alzheimer’s meta-analysis outcome population, we only know this for the PGC cohort. Therefore, we assumed that for the SNPs tested for in the ADSP and IGAP cohorts, every SNP had the same case and control number as the overall participant numbers. For ADSP 4,343 cases and 3,165 controls, for IGAP 17,008 cases and 14,471 controls. A prevalence of the outcome as required for Steiger filtering, we stated that the prevalence of the outcome was 0.07. Similarly, we do not know the exact case and control number for each SNP in the COPD exposure. Therefore, we assumed that every SNP had the same case and control number as the total number of case (35,735) and control (222,076) participants in the study. We estimated the prevalence to be 0.1 as per the discovery GWAS. Lung function is a continuous trait, so does not require estimation of prevalence to perform Steiger filtering.

## Appendix 3. Details of Sample Populations

**Table E1.**
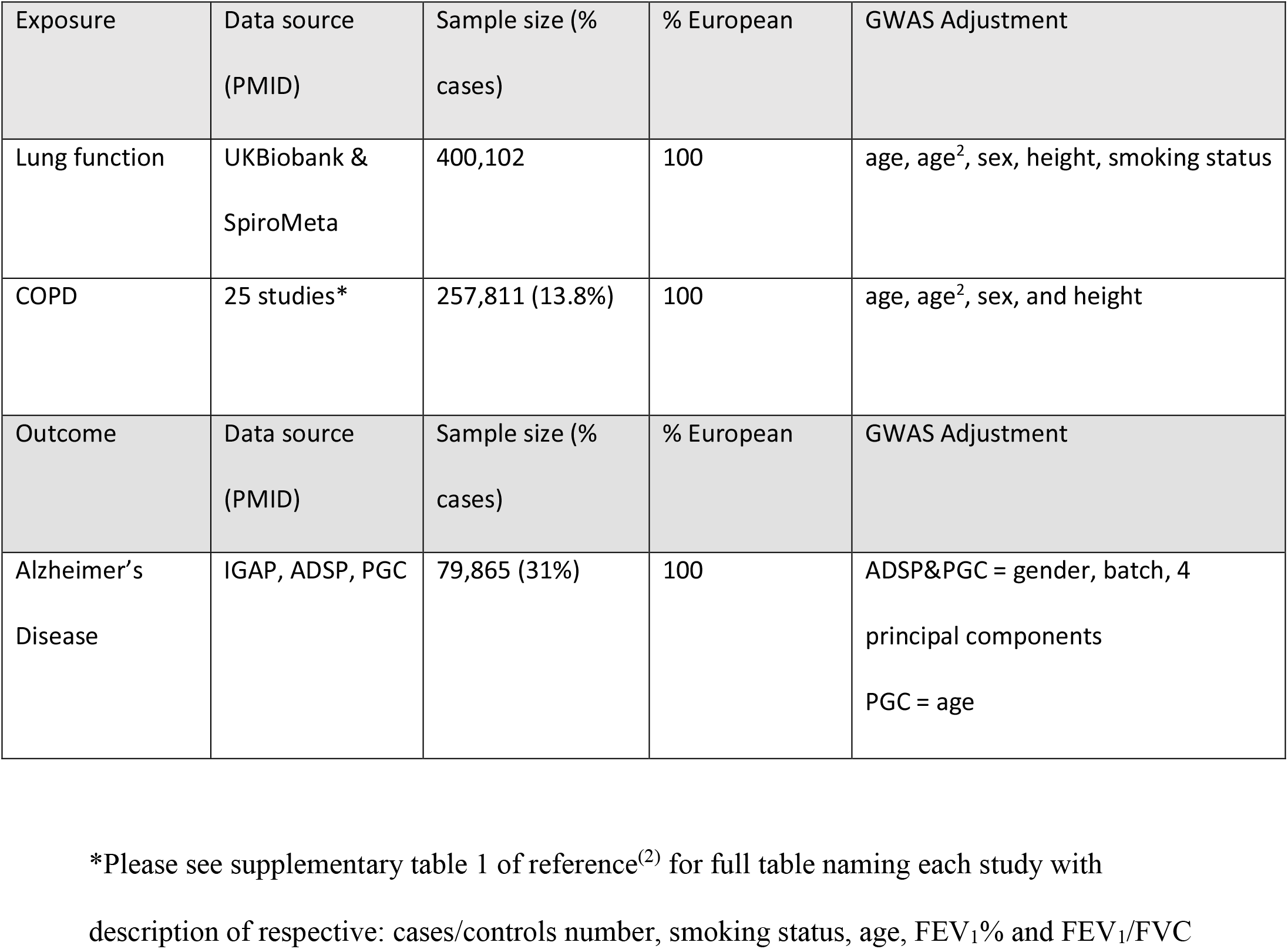
Description of GWAS samples used.

| Exposure | Data source<br>(PMID) | Sample size (%<br>cases) | % European | GWAS Adjustment |
| --- | --- | --- | --- | --- |
| Lung function | UKBiobank &<br>SpiroMeta | 400,102 | 100 | age, age<sup>2</sup>, sex, height, smoking status |
| COPD | 25 studies* | 257,811 (13.8%) | 100 | age, age<sup>2</sup>, sex, and height |
| Outcome | Data source<br>(PMID) | Sample size (%<br>cases) | % European | GWAS Adjustment |
| Alzheimer's<br>Disease | IGAP, ADSP, PGC | 79,865 (31%) | 100 | ADSP&PGC = gender, batch, 4<br>principal components<br>PGC = age |

\*Please see supplementary table 1 of reference<sup>(2)</sup> for full table naming each study with description of respective: cases/controls number, smoking status, age, FEV<sub>1</sub>% and FEV<sub>1</sub>/FVC

## Appendix 4. Flow chart of analysis

Study protocols were not pre-registered.

**Figure E5.**
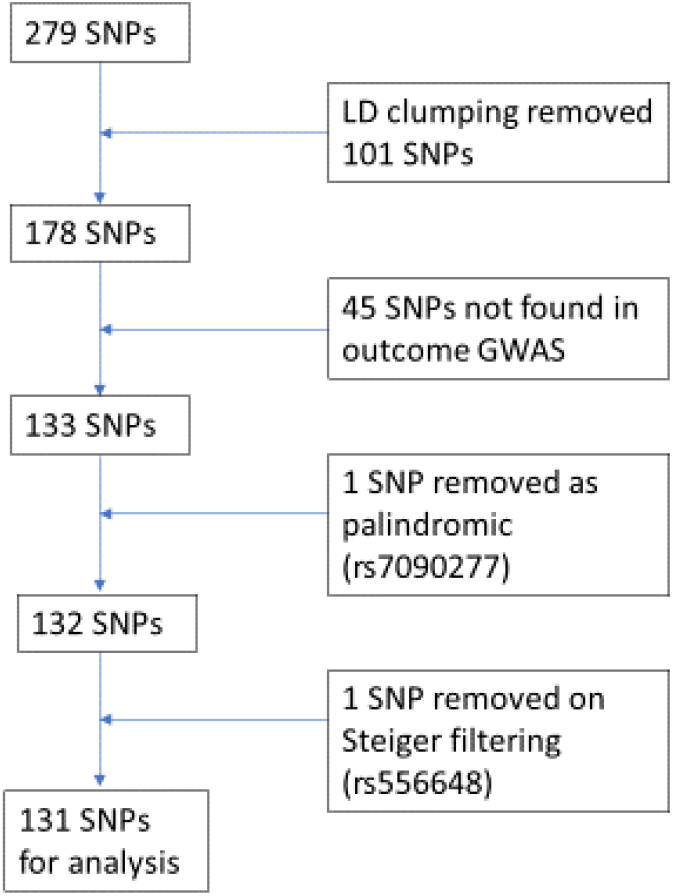
Flow chart of analysis for all lung function trait SNPs^(1)^

**Figure E6.**
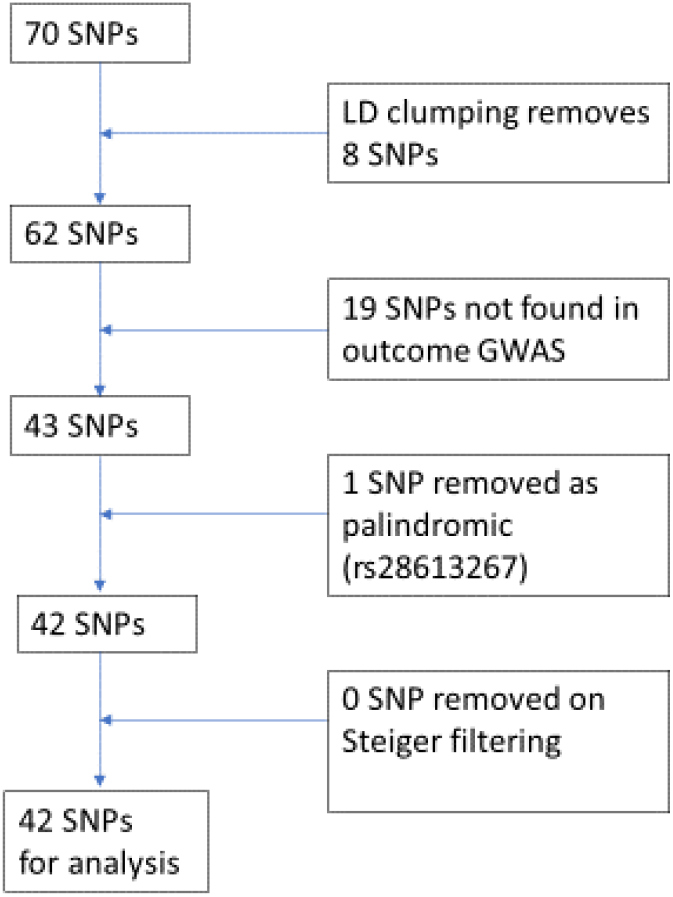
Flow chart of analysis for FEV_1_ SNPs.

**Figure E7.**
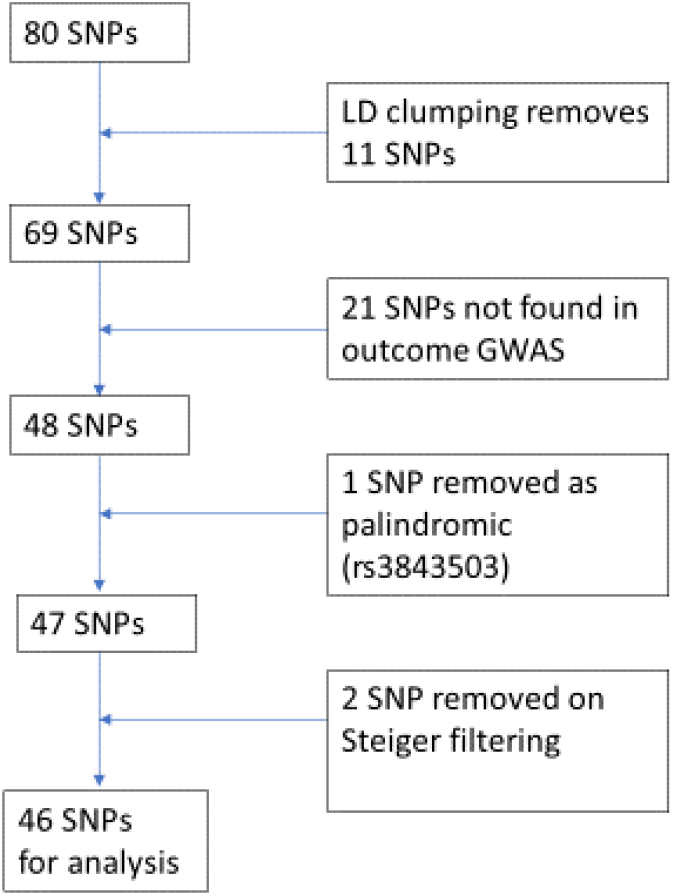
Flow Chart of analysis for FVC SNP’s.

**Figure E8.**
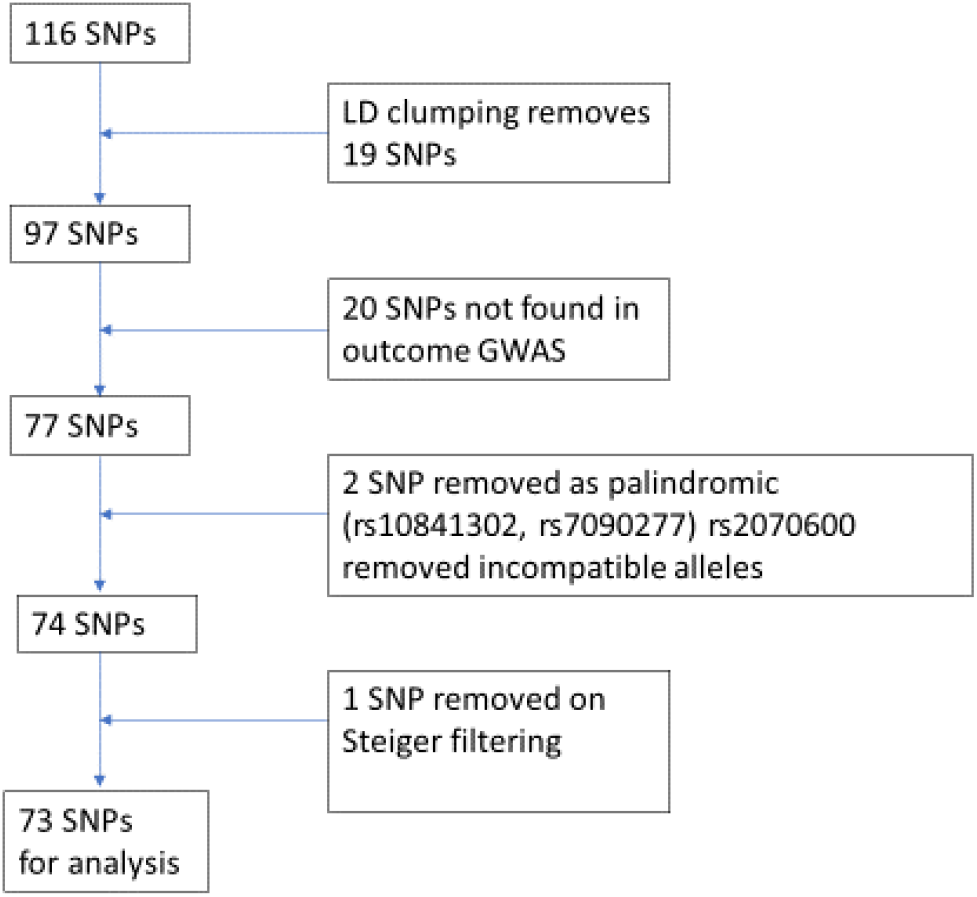
Flow Chart of analysis for effect FEV_1_/FVC SNPs.

**Figure E9.**
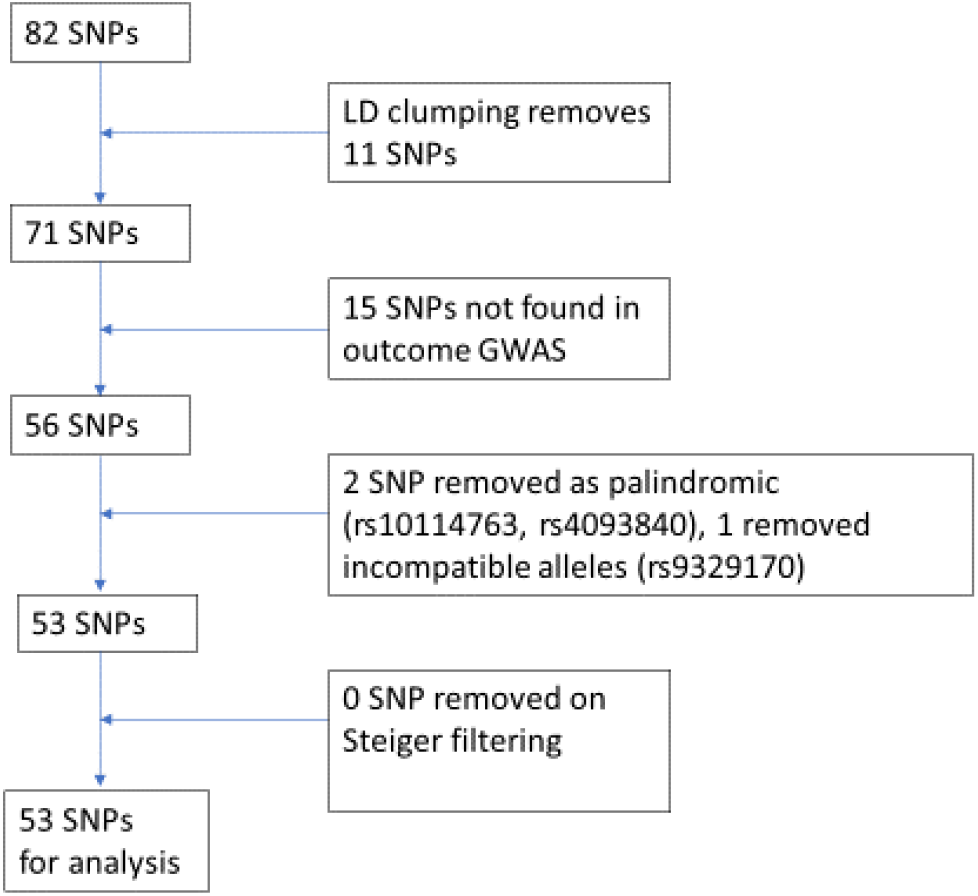
Flow Chart of analysis for COPD liability SNPs^(2)^

## Appendix 5

Code available on request from corresponding author.

Data used was summary data freely available in supplementary tables or from corresponding authors of GWAS.

